# Independent assessment of a deep learning system for lymph node metastasis detection on the Augmented Reality Microscope

**DOI:** 10.1101/2022.05.24.22275431

**Authors:** David Jin, Joseph H. Rosenthal, Elaine E. Thompson, Jared Dunnmon, Niels H. Olson

## Abstract

Several machine learning algorithms have demonstrated high predictive capability in the identification of cancer within digitized pathology slides. The Augmented Reality Microscope (ARM) has allowed these algorithms to be seamlessly integrated within the current pathology workflow by overlaying their inferences onto its microscopic field of view in real time. In this paper, we present an independent assessment of the LYmph Node Assistant (LYNA) models, state-of-the-art algorithms for the identification of breast cancer metastases in lymph node biopsies which have been optimized for usage at different ARM magnifications. We assessed the models on a set of 40 whole slide images at the commonly used objective magnifications of 10x, 20x, and 40x. We analyzed the performance of the models across clinically relevant subclasses of tissue, including metastatic breast cancer, lymphocytes, histiocytes, veins, and fat. We also analyzed the models’ performance on potential types of contaminant tissue such as endometrial carcinoma and papillary thyroid cancer. Each model obtained overall AUC values of approximately 0.98, accuracy values of approximately 0.94, and sensitivity values above 0.88 at classifying small regions of a field of view as benign or cancerous. Across tissue subclasses, the models performed most accurately on fat and blood, and least accurately on histiocytes, germinal centers, and sinus. The models also struggled with the identification of isolated tumor cells, especially at lower magnifications. After testing, we manually reviewed the discrepancies between model predictions and ground truth in order to understand the causes of error. We introduce a distinction between *proper* and *improper* ground truth to allow for analysis in cases of uncertain annotations or on tasks with low inter-rater reliability. Taken together, these methods comprise a novel approach for exploratory model analysis over complex anatomic pathology data in which precise ground truth is difficult to establish.

## Introduction

The evaluation of lymph nodes is a critical component of the breast cancer staging process which heavily informs treatment decisions (1, 2). The current diagnostic workflow, which involves microscopic review of the biopsy sample by a certified pathologist, is both time-consuming and error prone. Additionally, manual pathologist review of slides has been shown to exhibit poor inter-observer and intra-observer reliability (3, 4). While the re-examination of slides and other techniques such as immunohistochemical staining can reduce the probability of misdiagnosis (5), access to qualified experts to perform such analysis is often extremely limited (6). These shortages have been exacerbated by the COVID-19 pandemic, which has put a strain on medical laboratories all across the world (7, 8). A particularly stark example of these trends occurs in the Military Health System, which is experiencing a policy-driven decline in pathologist staffing on top of the existing shortage (9, 10).

Artificial intelligence may have the potential to address the limited availability of medical personnel. Indeed, recent advances in the application of deep learning-based computer vision techniques to digital pathology data have enabled the creation of algorithms that have demonstrated high predictive capability in the detection of breast and prostate cancer within whole slide images (WSI) (11, 12, 13, 14). In particular, the diagnostic accuracy of several deep learning algorithms designed to detect metastatic breast cancer within lymph node biopsy WSIs has been found to be comparable to that of pathologists without time constraint (15). Additionally, algorithm-assisted pathologists have been found to have higher accuracy, sensitivity, and speed than unassisted pathologists in the identification of micrometastases on digitized slides of lymph node biopsies (16).

These computer vision algorithms have been incorporated into the pathologist microscopy workflow by integrating microdisplays into traditional light microscopes to produce Augmented Reality Microscopes (ARMs). The ARM superimposes real-time inferences from a machine learning algorithm onto the current microscopic field of view (FOV), providing its inference to the pathologist as decision support. Combined with an algorithm with high predictive performance, the ARM has the potential to increase both diagnostic accuracy and efficiency (17).

Various deep learning models that have demonstrated success in the diagnosis of WSIs have been modified for use on the ARM. One such algorithm is the LYmph Node Assistant (LYNA) model, which is a state-of-the-art algorithm for detecting cancer metastases within sentinel lymph node biopsies (18). On the ARM platform, the LYNA models take a microscopic FOV at a given resolution, divide it into small regions, and output a cancer-likelihood prediction for each region within that FOV. These predictions are then presented as outlines, superimposed onto the microscopic FOV, that indicate the cancerous regions to the viewing pathologist (17). Similar to immunohistochemistry analysis, these algorithms have the potential to become highly effective decision support tools. However, unlike immunohistochemistry, where issues have been identified (e.g. cross-reactants and interfering substances) and mitigation strategies have been developed (e.g. peroxidase blocking), the limitations of these algorithms have not been quantified or mitigated. While these algorithms have demonstrated high predictive performance on WSIs, the utility of an algorithm on the ARM depends on its ability to delimit particular areas of cancer within an FOV. The performance of an algorithm on this segmentation task requires far greater precision than the classification of WSIs and has not been extensively studied in the literature. Additionally, while machine learning algorithms have shown promising performance on cancer classification, we do not have a detailed understanding of the robustness or consistency of their performance across the clinically relevant subclasses of tissue. This gap is significant because machine learning models for medical imaging commonly suffer from hidden stratification, i.e., highly variable performance across the clinically relevant subclasses (19). In this paper, we present an independent assessment of the LYNA models on a test set of FOVs derived from 20 benign and 20 cancer-containing WSIs. In order to understand the models’ utility for decision support, we measured their ability to detect cancer within small regions of an input FOV. We present detailed metrics of the models’ overall performance, as well as their performance on operationally realistic subclasses of tissue within the lymph node, including metastatic breast cancer, lymphocytes, histiocytes, germinal centers, veins, arteries, and fat. We found that the models performed best on fat and struggled most on histiocytes, which is consistent with the experience of pathologists.

Drawing annotations with sufficient fidelity to evaluate performance on these small regions is both time-consuming and extremely difficult, since tumor and tissue boundaries may be too complex to reasonably delineate. These challenges compound upon the already poor inter-observer reliability between pathologists (3, 4). In order to analyze this and other cases of uncertain ground truth, we introduce the concept of *proper* and *improper* ground truth annotations. Using this distinction, we analyze both model performance and ground truth as a source of potential error. Our analysis of ground truth uncovered common classes of annotation errors, such as the over-labeling of cancer around complex tumor margins, which should be noted for future studies.

## Materials and Methods

### Model details

We tested three versions of the LYNA model which were optimized for the commonly used objective microscope magnifications of 10x, 20x, and 40x. Each model was trained to perform inference on FOVs at its corresponding magnification. These models are ARM-specific adaptations of a deep learning model that operates on WSIs (17). Testing of the LYNA model before its adaptation into the ARM modality can be found in Steiner *et al*. (2018) and Liu, *et al*. (2019) (16, 18).

Each of the LYNA models takes a microscopic FOV of the appropriate magnification and subdivides the FOV into a 14 × 14 grid of smaller squares. Each square, which we will refer to as a *region of interest (ROI)*, is 128 × 128 pixels in size and makes up 1/196th of the area of an FOV. An ROI is analogous to a “patch” from Chen *et al*. (2019) and Liu *et al*. (2019) (17, 18). The model performs inference on each ROI in a sliding-window fashion, taking context from the neighboring ROIs to inform its predictions. Ultimately, the model outputs a two-dimensional matrix of cancer predictions for each input FOV, with each entry of the matrix corresponding to an ROI. On the ARM, this matrix of cancer predictions is used to build a semantic segmentation of the FOV into cancerous and benign regions which is superimposed on the microscopic view. For each ROI within an FOV, the output cancer prediction is a value from 0.0 to 1.0, with 0.0 representing the lowest confidence that an ROI contains cancer, and 1.0 representing the highest confidence that an ROI contains cancer. This value is transformed into a binary classification of “cancer” or “benign” using the default cancer threshold of 0.5. In other words, an ROI with an output prediction of 0.5 or greater is classified as “cancer”, while an ROI with an output prediction of less than 0.5 is classified as “benign”. This is the threshold that the model operates by when deployed in the ARM system. Figure 1 presents an example FOV, its constituent ROIs, and the model’s predictions for each ROI.

**Figure 1.**
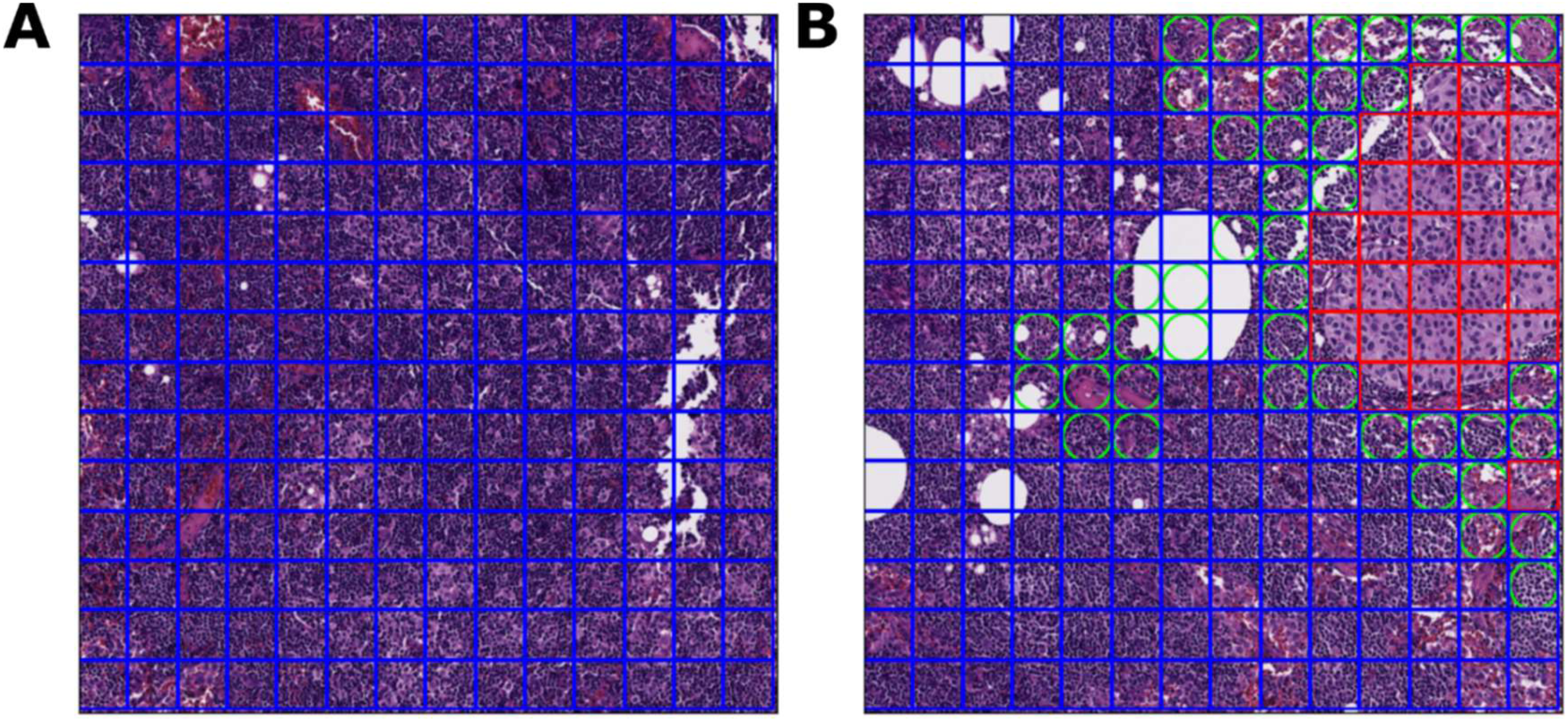
ROI and FOV examples at 20x magnification. **1A**. Within this FOV, each box designates an ROI. The blue border of each box indicates that the bounded ROI is ground truth “benign”. **1B**. Within this FOV, each red box indicates an ROI with a ground truth of “cancer”. A green circle within an ROI represents an area where the model’s classification diverged from the ground truth. Thus, an ROI with a green circle inscribed in a blue box is a false positive, while an ROI with a green circle inscribed in a red box is a false negative.

### Test data

We consider two test sets within this paper: the primary test set and the out-of-domain test set. Unless otherwise indicated, results correspond to the primary test set.

The primary test set consisted of 40 WSIs of lymph node sections from breast cancer patients who underwent a mastectomy or a lumpectomy with sentinel node biopsy. These WSIs were digitized with an Aperio AT2 scanner with a pixel size of 0.252 × 0.252 μm^2^. Of the 40 WSIs, 20 contained metastatic breast cancer and 20 were entirely benign. Macrometastases, micrometastases, and isolated tumor cells (ITCs) were all represented in the 20 cancerous WSIs. Each WSI was selected at random within its class (cancerous or benign) from separate cases at Naval Medical Center San Diego.

A single board-certified anatomic pathologist digitally labeled all 40 WSIs. Pan-cytokeratin immunostains were used to aid the pathologist labeling. These immunostains were produced by destaining and restaining the previously digitized H&E slide. In the labeling process, each slide was exhaustively annotated by outlining regions of each of the following clinically relevant subclasses of tissue: metastatic breast cancer (BrCA), histiocytes, germinal center (GC), mantle, lymphocytes, fat, sinus, capsule, nerve, artery, vein, and blood. Each tissue subclass was associated with its respective SNOMED code. This level of annotation may be compared to the 10 WSIs labeled with “detailed lesion annotations” in Bandi, *et al*. (2018) and the 159 WSIs with detailed metastases labeled in the Benjordi, *et al*. (2017), but with other tissue subclasses similarly annotated within our test set (20, 15).

Due to practical time constraints, the labeling pathologist was restricted to four hours of annotation time per slide. In comparison, the time-constrained pathologists annotating the CAMELYON16 dataset were given two hours to label a glass slide on a five-point scale ranging from “definitely normal” to “definitely cancer”, a comparatively simpler labeling process (15). Our highly detailed labeling process enabled the analysis of model performance across tissue subclasses but also produced some mislabeled data. We later analyzed the dataset for potential errors in ground truth annotations.

For a given magnification (10x, 20x, or 40x), we obtained FOVs by partitioning the WSIs into 1800 × 1800-pixel squares, matching the magnification and pixel size of the corresponding microscopic FOV. We partitioned WSIs to emulate microscopic FOVs in order to produce a large amount of data with cancerous regions and other tissue types outlined and annotated. This approach is not without limitations. In particular, our test FOVs lacked microscope artifacts such as vignetting and fisheye which do exist when observing slides on the ARM (21). For each FOV, each ROI was given a subclass and ground truth label with the following procedure. ROIs containing breast cancer were given the subclass label “BrCA”. Other ROIs were given their subclass label based on the primary tissue type within their region. These subclasses were then rolled up into ground truth annotations of “benign” or “cancer”. The ROIs in subclass “BrCA” were classified as “cancer”, while the ROIs in other subclasses were classified as “benign”. ROIs whose region was unlabeled (primarily due to whitespace) were not mapped to a ground truth annotation and thus not included in the test set. In total, the primary test set included 358,285 ROIs at 10x magnification, 1,448,284 ROIs at 20x magnification, and 5,802,458 ROIs at 40x magnification.

Each FOV was also given a ground truth annotation. An FOV was assigned ground truth “cancer” if any of its constituent ROIs were ground truth “cancer” and was assigned ground truth “benign” otherwise. In total, the primary test set included 2,905 FOVs at 10x magnification, 10,018 FOVs at 20x magnification, and 35,554 FOVs at 40x magnification.

The out-of-domain test set consisted of common types of contaminant tissue (“floaters”), including papillary thyroid cancer, papillary urothelial carcinoma, endometrial carcinoma, embryonal carcinoma, high-grade carcinoma, and serous borderline tumor. For each of the six aforementioned tissue types, a representative WSI was selected by the labeling pathologist. The cancer tissue within each WSI was outlined and annotated by the same pathologist. After annotation, the WSIs were partitioned into FOVs at 10x magnification and then further partitioned into ROIs. The out-of-domain test set was the subset of these ROIs which contained cancer tissue. More information about the test sets, data labeling procedure, and FOV creation procedure can be found in the supplementary materials.

### Test methodology

The testing procedures for the 10x, 20x, and 40x LYNA models were performed independently from each other using the test sets of corresponding resolutions. Testing of the LYNA models on the primary and the out-of-domain test sets was also performed separately.

For the primary test set, we computed performance metrics over the set of ROIs and over the set of FOVs. Since the LYNA models do not return a classification for an FOV, the model classification for each FOV was obtained by aggregating the values of its constituent ROIs. An FOV was classified as “cancer” if any of its ROIs were classified as “cancer”, otherwise it was classified as “benign”. This aggregation procedure is the same as the one used by Chen *et al*. (2019) (17).

For the ROI and FOV-level tasks, we computed the accuracy, sensitivity, specificity, positive predictive value (PPV), and negative predictive value (NPV) for each model across its corresponding test set. We also generated receiver operating characteristic (ROC) curves and computed area under the curve (AUC) values. Within the ROI-level task, we computed the same metrics for each model across each of the clinically relevant subclasses. We also performed an ablation study, removing the extremely common and distinguishable ‘Fat’ subclass in order to demonstrate its impact on average performance metrics. For the out-of-domain test set, we computed performance metrics at the ROI level at 10x magnification. All ROIs within the test set were cancerous, so we computed the proportion of ROIs that each model correctly classified as cancer over the entire test set and over each type of cancer.

### Re-review methodology

After computing the models’ performance over the primary test set, we manually reviewed the ROIs where the models’ predictions diverged from the ground truth. We considered not only the models’ predictions, but also the ground truth as a potential source of error. The complexities of tumor boundaries, time constraints on labeling, and the difficulty of detailed ROI-level annotations (especially in the case of scattered tumor cells) all may contribute to potential error in the ground truth labels. This is not out of the ordinary, as individual pathologists often disagree about which regions on a slide are cancerous (3).

Due to the difficulty of accurately labeling ROI-level ground truth, we introduce the concept of *proper* and *improper* ground truth annotations. In a *proper annotation*, ground truth is judged to be correct on detailed re-review. In an *improper annotation*, the ground truth is judged to be incorrect on detailed re-review. For instance, a *proper false negative* is an ROI that the model classified as negative, is ground truth positive, and on re-review, the ground truth is judged to be correct. On the other hand, an *improper false negative* is an ROI that the model classified as negative, is ground truth positive, but on re-review, the ground truth is judged to be incorrect. The [proper / improper] distinction adds another dimension to the traditional [true / false] - [positive / negative] paradigm. It is particularly useful to describe results obtained with uncertain ground truth and can help to characterize the performance of an annotator. For instance, a tissue subclass found to have a large percentage of improper false positives may indicate a subclass for which the annotator is underperforming, and the AI model may provide value. Due to the labeling challenges mentioned above, a proportion of the models’ errors are improper. In order to classify an ROI’s annotation as proper or improper, the original labeling pathologist re-compared the ROI to the corresponding area on the slide’s immunostain. We empirically selected a subset of the false negative containing FOVs for re-review. Within the Examination of errors section, we present a number of illustrative examples and characterize each as proper or improper.

### Software and hardware

Model inference was performed using the Themis test harness developed by MORSE Corp to support testing and evaluation at the Department of Defense Joint Artificial Intelligence Center. Themis is designed to take in models and perform inference on test sets in a way that is repeatable and standardized in order to compare models of different versions or from different developers.

The partitioning of each WSI into FOVs and the generation of ground truth classifications for each ROI was performed using the Norm software library developed at The Henry M. Jackson Foundation for the Advancement of Military Medicine. Norm enables the rapid conversion of WSIs to annotation-associated FOVs from both XML and protobuf-specified annotations in a highly parallelized fashion, using OpenSlide 3.4.1 for SVS slide processing and OpenCV 3.2 for image processing.

Model inference was performed on a Google Cloud Platform instance running Debian 10 with 120GB of memory and 4 NVIDIA K80 GPUs. Debian 10 was selected due to limitations of an OpenSlide dependency. During testing, the GPUs were run in parallel, each running a single inference task independently from the others. In comparison, the ARM is powered by a single NVIDIA Titan Xp (17). The NVIDIA K80 contains 4,992 CUDA cores, while the NVIDIA Titan Xp contains 3,840 CUDA cores.

## Results

### ROI-level results

We evaluated each model on the corresponding test set and computed the results at the ROI level. The performance of an ARM model on these small ROIs is crucial for its ability to accurately outline cancer and function as a decision support tool for pathologists.

The results over the entire set of ROIs for each resolution are presented in Table 1. The ROC curves for each resolution are presented in Figure 2A. All models attained accuracy values of approximately 0.94 and AUC values of approximately 0.98. Across the models, specificity increased slightly with magnification while sensitivity decreased. Each model yielded an NPV of over 0.98 and a PPV of over 0.6.

**Table 1.** ROI and FOV level results summary

| Table 1. ROI and FOV level results summary |  |  |  |  |  |  |  |
| --- | --- | --- | --- | --- | --- | --- | --- |
| Model | Mode | AUC | Accuracy | Sensitivity | Specificity | PPV | NPV |
| LYNA 10x | ROI | 0.981 | 0.938 | 0.930 | 0.938 | 0.604 | 0.993 |
|  | FOV | 0.938 | 0.746 | 0.939 | 0.716 | 0.342 | 0.987 |
| LYNA 20x | ROI | 0.978 | 0.939 | 0.897 | 0.943 | 0.601 | 0.990 |
|  | FOV | 0.959 | 0.792 | 0.956 | 0.771 | 0.347 | 0.993 |
| LYNA 40x | ROI | 0.976 | 0.947 | 0.878 | 0.954 | 0.641 | 0.988 |
|  | FOV | 0.948 | 0.823 | 0.948 | 0.809 | 0.354 | 0.993 |

**Figure 2.**
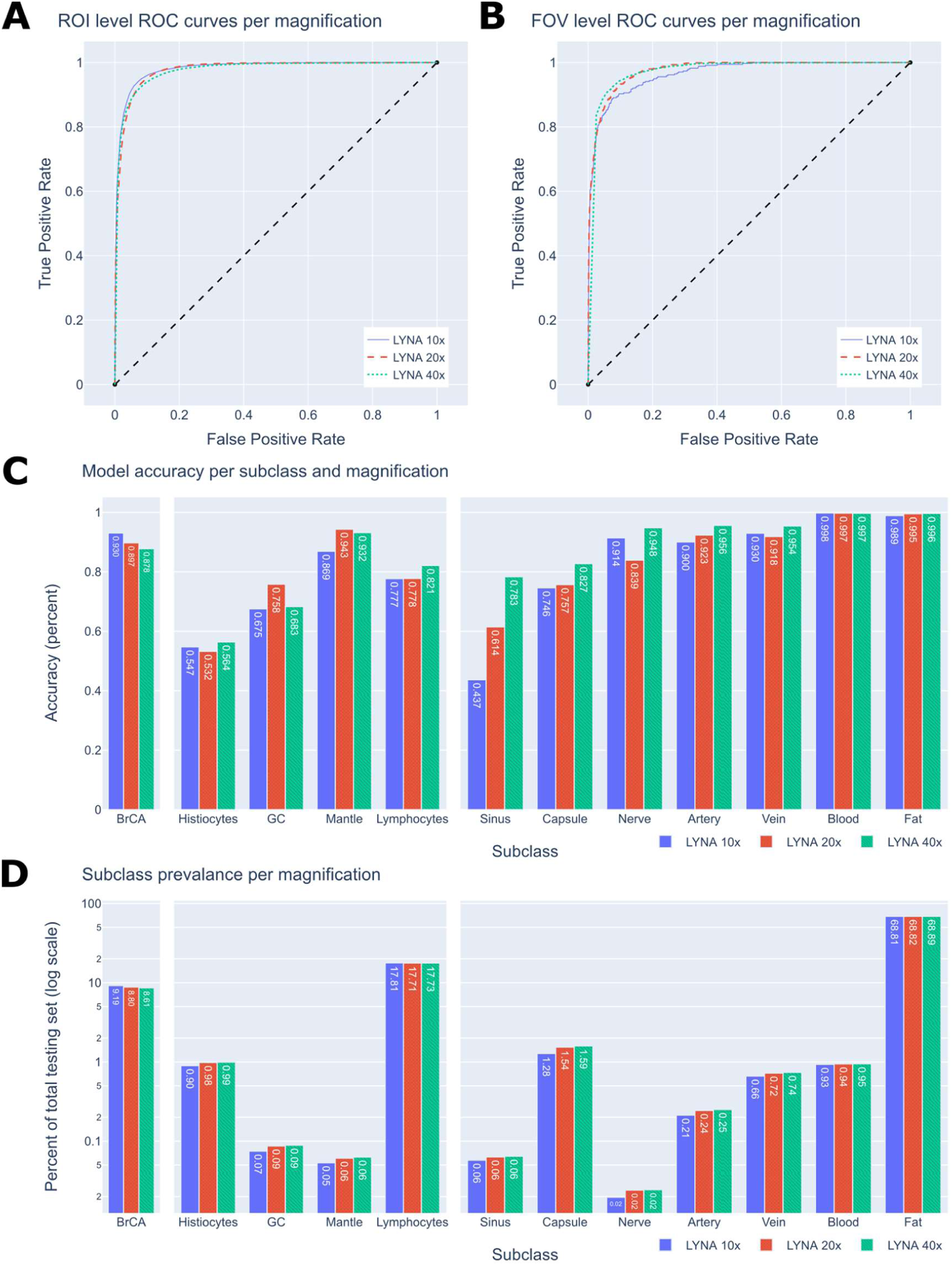
ROC curves and subclass performance graphs. **2A**. ROI-level ROC curves. **2B**. FOV-level ROC curves. **2C**. The accuracies of each subclass within the 10x, 20x and 40x datasets. The subclasses are divided into the three categories of cancer, immune cells, and connective tissue. **2D**. The prevalence of each subclass within the 10x, 20x, and 40x datasets.

We also computed each models’ results across the clinically relevant tissue subclasses. The models’ results across each subclass are presented in Table 2 and Figure 2C. The prevalence of each subclass within the test set is presented in Table 2 and Figure 2D. All models performed best on fat and blood, achieving accuracies of approximately 0.99. All models struggled similarly with histiocytes, likely due to their visual similarity with cancer, achieving accuracies of approximately 0.55. When restricted to ROIs of the metastatic breast cancer subclass, the models’ performance decreased with magnification, attaining an accuracy of 0.93, 0.90, and 0.88 at the 10x, 20x, and 40x magnifications, respectively. However, the 40x model performed better than the 10x and 20x models for most of the benign subclasses, with the exception of germinal center and mantle zone. Performance on sinus tissue drastically improved with increasing magnification, with accuracy rising from 0.44 at 10x to 0.61 at 20x and finally to 0.78 at 40x magnification. For each metric, we computed a 95% confidence interval which is presented in the supplementary materials.

**Table 2.**
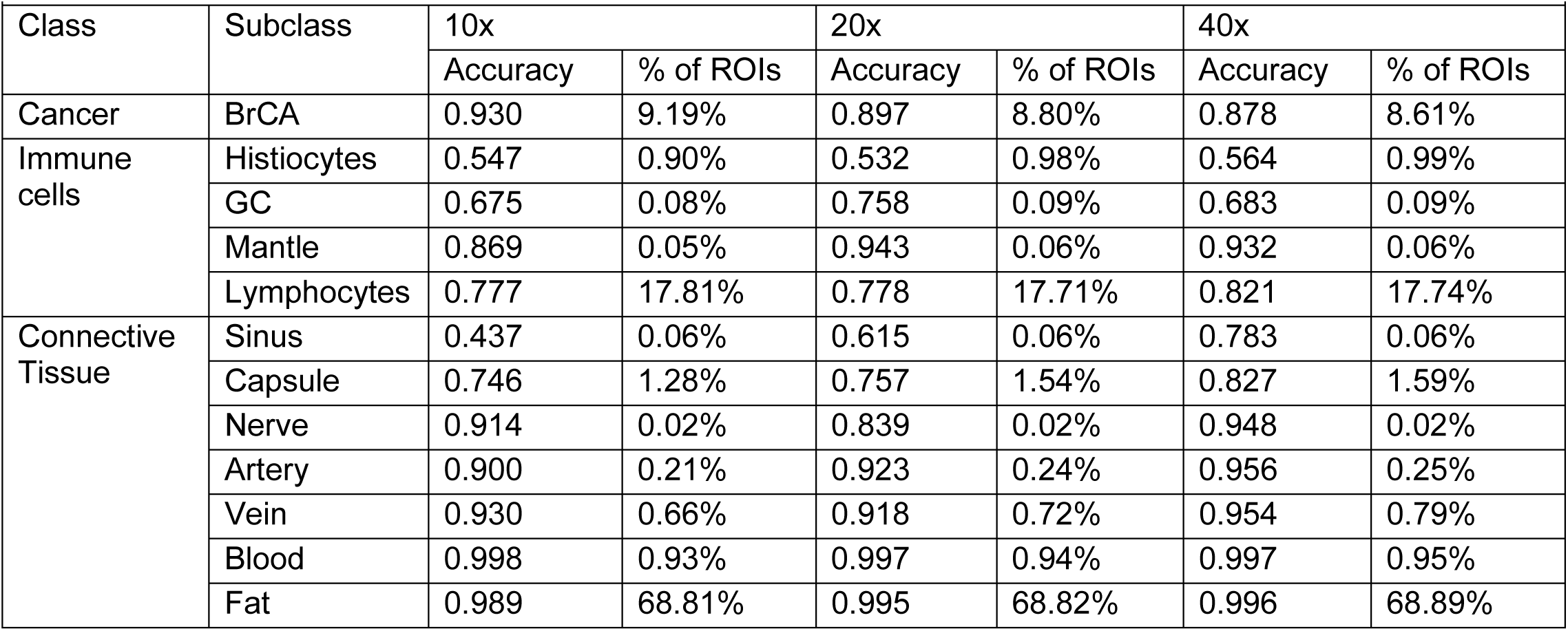
Results per subclass per magnification

We also considered the performance of each model after removing the “fat” subclass from the test set. The fat subclass was the most prevalent subclass by far, making up approximately 69% of ROIs at each magnification, and it was also a subclass on which each model performed extremely well. The removal of the fat subclass dropped the AUC, accuracy, specificity, and NPV significantly compared to the overall test sets. For example, the AUC values decreased from 0.98 to 0.94 at 10x magnification, 0.98 to 0.92 at 20x magnification, and 0.98 to 0.92 at 40x magnification. For each model, the PPV without fat was slightly increased from the overall test sets due to the removal of over 70% of the negative instances. A detailed comparison of the results on the overall ROI test set and the test set without the fat subclass is presented in the supplementary materials.

### FOV-level results

An algorithm does not need to correctly outline all the cancer on an FOV to be useful; it may suffice that it detects enough cancer to give the pathologist pause. Within this section, we present metrics regarding the model’s performance at the task of classifying an FOV as “cancer” or “benign”.

The results over the entire set of FOVs for each resolution are presented in Table 1. The ROC curves for each resolution are presented in Figure 2B. Across all magnifications, the FOV-level sensitivity and NPV metrics are comparable or better than their ROI-level counterparts. However, the models obtain worse performance across all other metrics. Notably, the models obtain low PPVs between 0.34 and 0.36.

### Out-of-domain performance

Contaminant tissue (also known as “floaters”) may appear on a microscope slide during slide preparation and may impact a patient’s diagnosis (22). In order to understand the performance of the LYNA model on this source of error, we studied the model’s behavior on several common tumor types known to be friable or otherwise problematic. In particular, we considered WSIs containing papillary thyroid cancer, papillary urothelial carcinoma, endometrial carcinoma, embryonal carcinoma, high-grade carcinoma, and serous borderline tumor. Figure 3 presents several example FOVs of the models’ inferences on these types of tissue.

**Figure 3.**
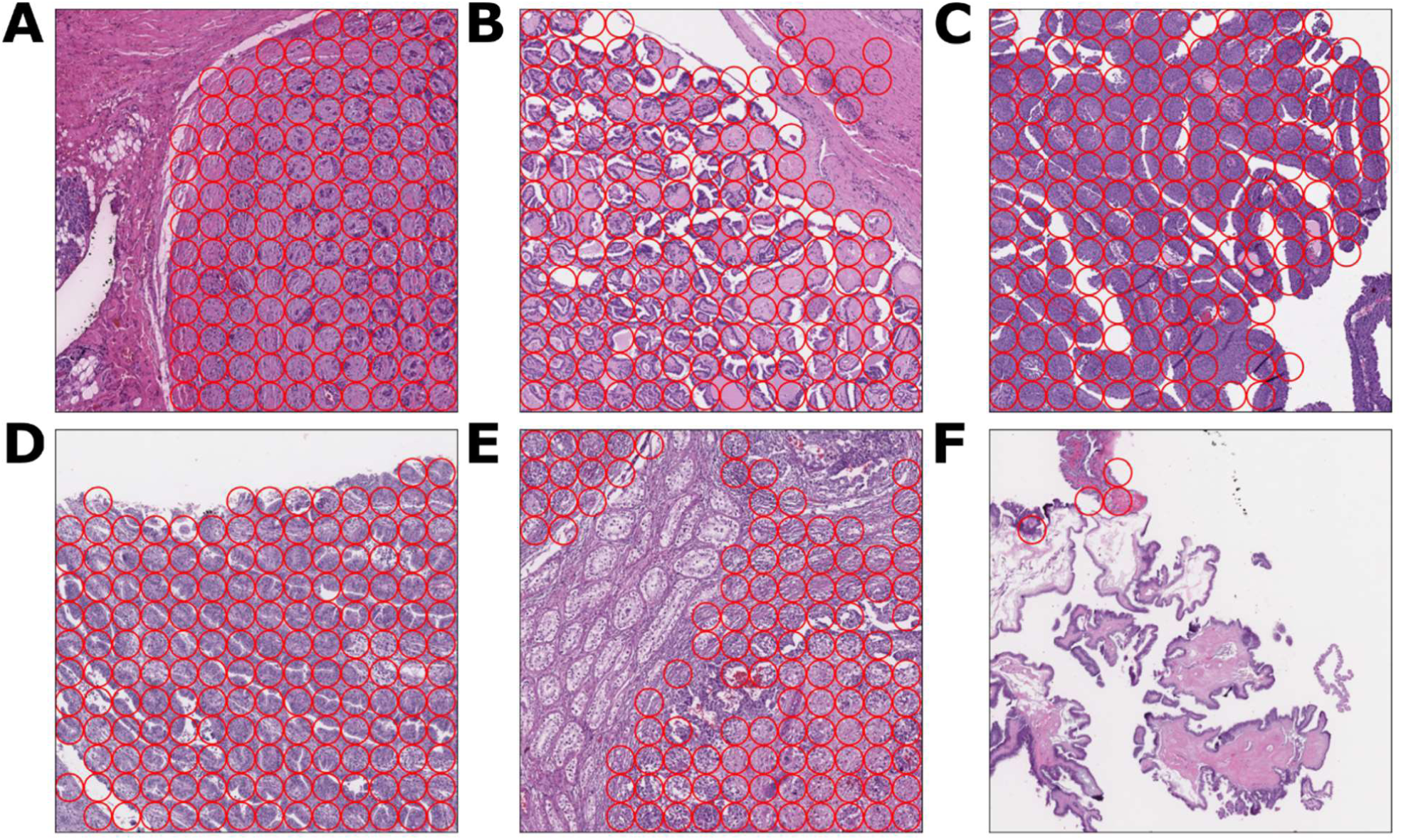
10x magnification FOVs of common out-of-domain tumor tissues. In each FOV, each ROI with a red circle indicates a region that the ROI classified as cancer. We use this indicator to differentiate out-of-domain ROIs from the metastatic breast cancer ROIs in the primary test set. **3A**. High-grade carcinoma FOV with model cancer classifications. **3B**. Papillary thyroid cancer FOV with model cancer classifications. **3C**. Papillary urothelial carcinoma FOV with model cancer classifications. **3D**. Endometrial carcinoma FOV with model cancer classifications. **3E**. Embryonal carcinoma FOV with model cancer classifications. **3F**. Serous borderline tumor FOV with model cancer classifications.

For each tissue type, we evaluated the model on the ROIs which were labeled as cancer at 10x magnification. The model obtained positive predictive values of 0.95 for high-grade carcinoma, 0.90 for papillary thyroid cancer, 0.73 for papillary urothelial carcinoma, 0.67 for embryonal carcinoma, 0.58 for endometrial carcinoma, and 0.09 for serous borderline tumor. More details about the performance on the out-of-domain test set is presented in the supplementary materials.

### Examination of errors

We present a number of illustrative examples of FOVs containing errors in order to give insight into the ARM models’ performance. We categorize each of these examples as proper or improper and explain the hypothesized cause behind the divergence of prediction and ground truth. See Materials and Methods for definitions of *proper* and *improper* ground truth. Within the context of the decision support task, the most impactful class of errors are false negatives since a model’s non-detection of a cancerous region may have far worse consequences than its wrongful detection of cancer. For this reason, we empirically selected a subset of the LYNA models’ false negative classifications in order to understand the underlying causes of these errors.

Proper and improper false negative classifications are presented in Figure 4A and 4C respectively. In the case of Figure 4A, the model missed a large region of cancer within the center of the FOV. These false negative classifications were confirmed to be proper by the immunostain in Figure 4B. In the case of Figure 4C, the ground truth annotation of cancer was drawn around the entire region of dark tissue. The labeling pathologist drew this annotation around this entire region at low magnification instead of the individual clusters of cancer, as correctly shown in the immunostain in Figure 4D. In this case, a higher-resolution annotation would have been more correct but would have also been significantly more difficult and time-consuming.

**Figure 4.**
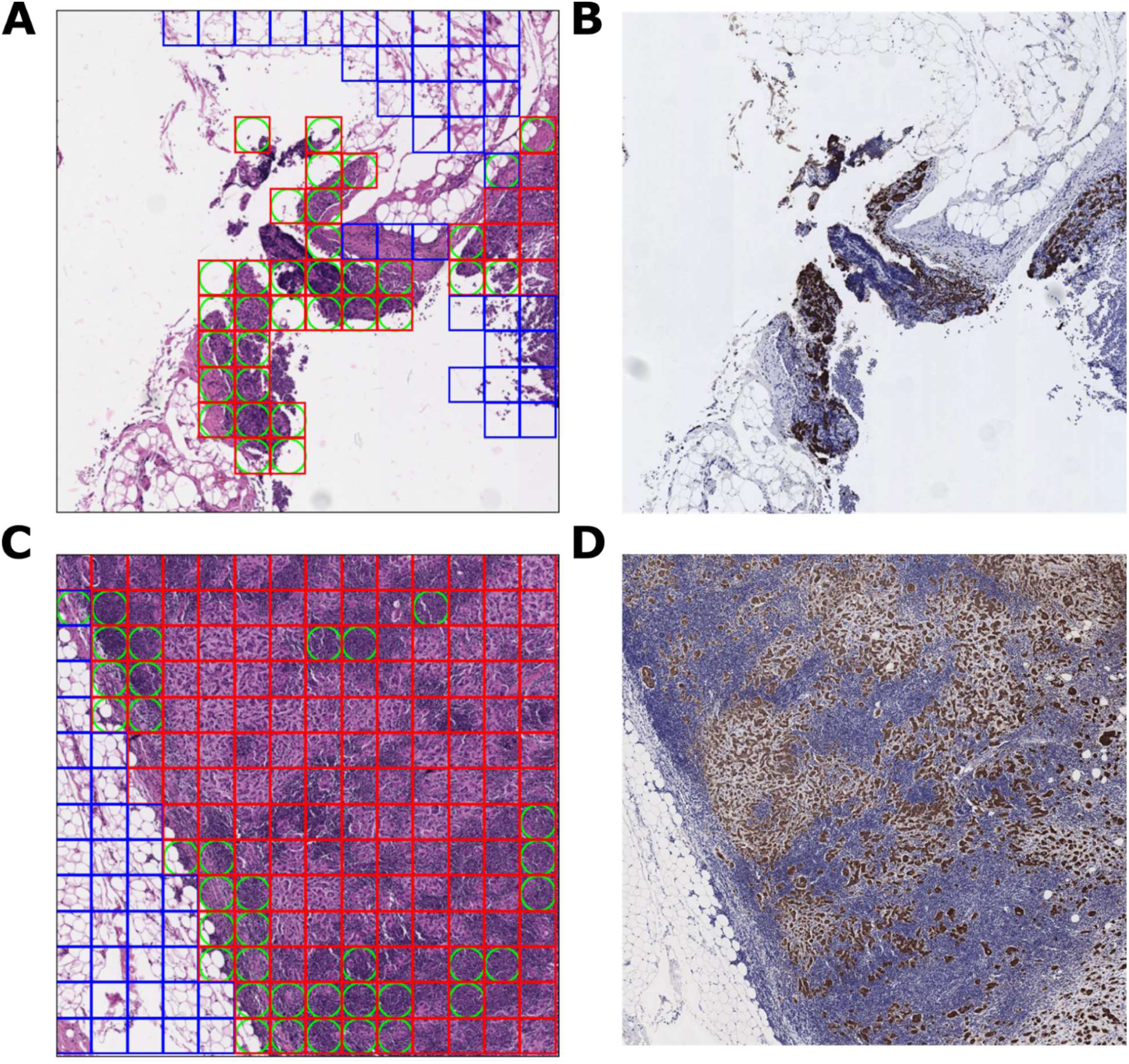
10x magnification FOVs with proper and improper false negatives. **4A**. A 10x FOV which contains numerous false negative ROIs, which are indicated by the red boxes (ground truth cancer) inscribed with green circles (machine disagreement with ground truth). **4B**. An immunostain of the FOV in 4A. Brown regions of the immunostain indicate cancer. The immunostain confirms the ground truth of the central region, thus the ROIs within this region are proper false negatives. **4C**. A 10x FOV which contains improper false negative ROIs due to the over-labelling of cancer by our annotating pathologist. On review, the annotating pathologist agreed that many of the above false negatives were improper. **4D**. An immunostain of the FOV in 4C. This immunostain confirms the over-labelling of cancer.

Another FOV containing proper false negative classifications is presented in Figure 5A. On re-review, the 20x FOV is uniformly cancerous, but obtains a wide range of ROI predictions. This behavior is limited to the 20x model; the 10x and 40x models confidently and accurately classify the corresponding regions as cancer. The behavior of the 40x LYNA model is shown in Figure 5B. The heatmaps in 5A and 5B illustrate the different ranges of ROI predictions on the two FOVs.

**Figure 5.**
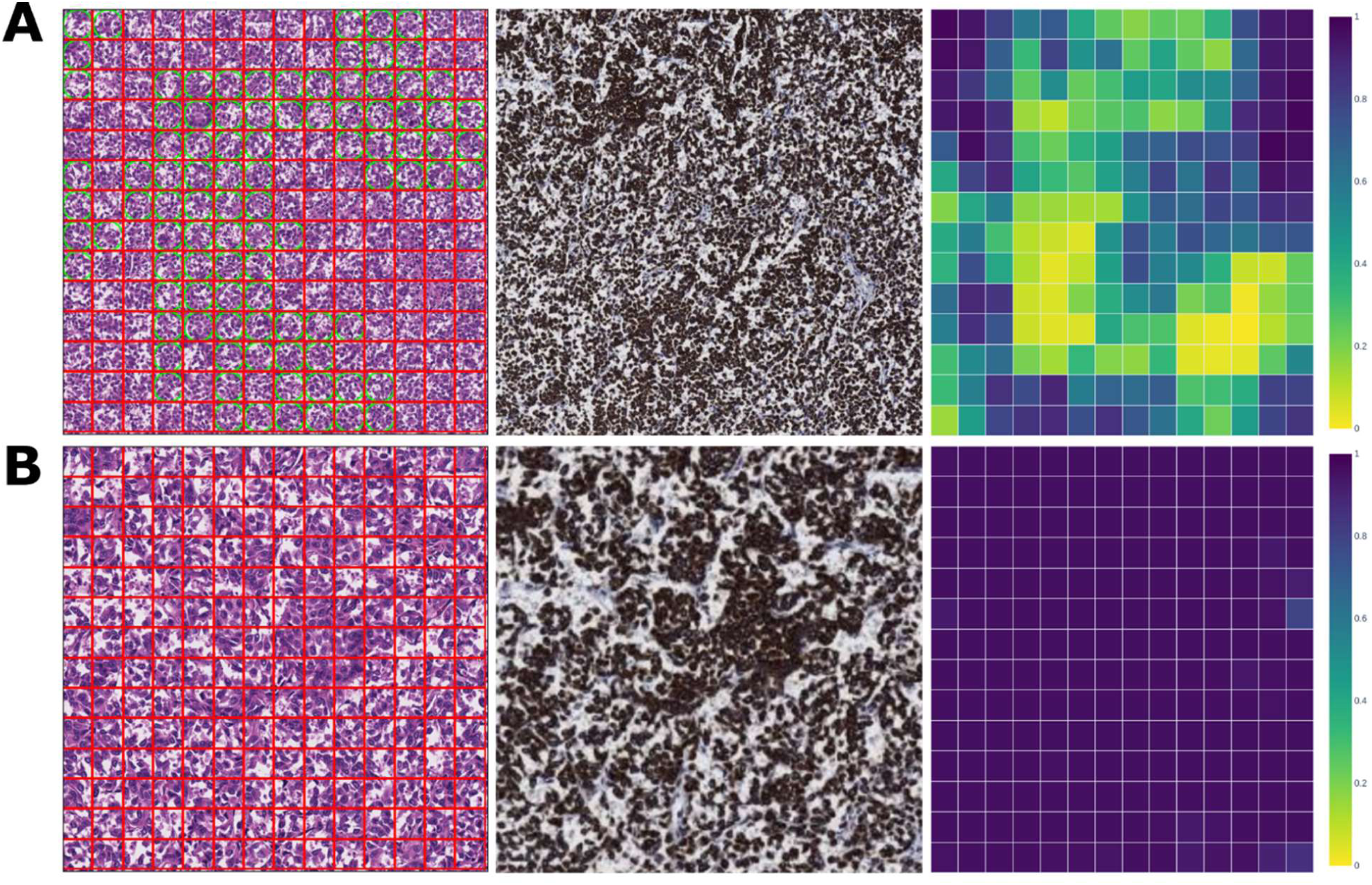
20x & 40x magnification FOVs with proper false negatives. **5A**. A 20x FOV with model predictions, immunostain, and prediction heatmap. This FOV contains a large number of proper false negatives. Despite the visual uniformity of this FOV, the set of ROI predictions are extremely varied, as indicated in the heatmap. **5B**. A 40x FOV with model predictions, immunostain, and prediction heatmap. This FOV is the upper left quadrant of the above 20x FOV. The ROIs in this FOV are confidently classified as cancer.

An illustrative example of an area containing proper false negatives of isolated tumor cells (ITC) is presented in Figures 6A and 6B at 20x and 40x magnifications respectively. In these FOVs, ITCs are the only tumor tissue within the view. The ROIs containing these ITCs are misclassified by the models at both magnifications. However, in the 20x magnification, many of the ITC-containing ROIs are mislabeled as benign in the ground truth. This can be seen by comparing the ROI ground truth with the immunostain in Figure 6A. These are examples of improper true positives, where the model’s classifications and ground truth agree, but ground truth is judged to be incorrect on re-review. More examples of errors and their analysis can be found in the supplementary materials.

**Figure 6.**
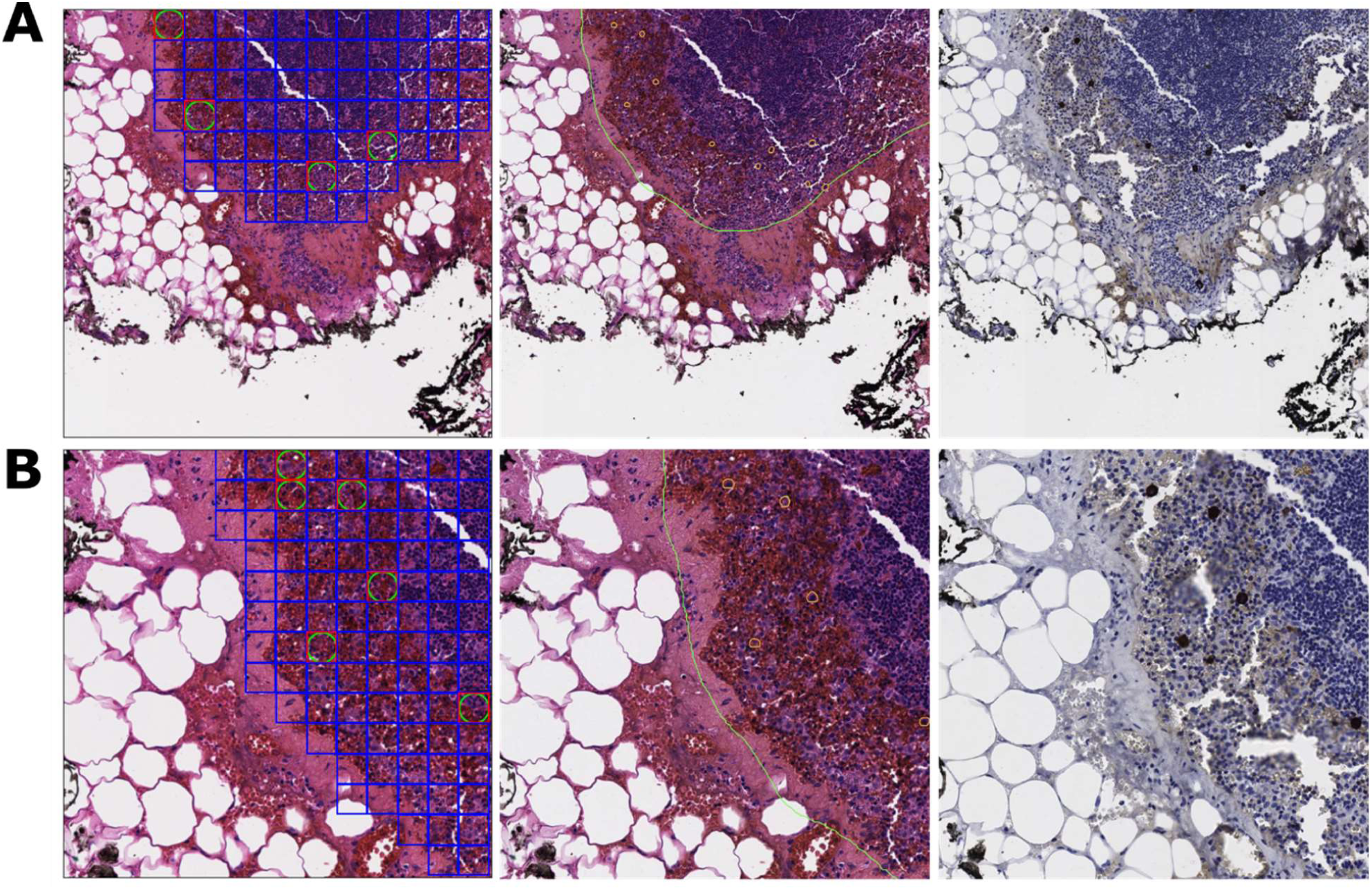
20x & 40x magnification FOVs with ITC proper false negatives. **6A**. A 20x FOV with model predictions, annotations, and immunostain. ITCs are the only tumor tissue in the FOV and are annotated with small orange circles in the annotations. All false negatives in this FOV are proper. Some ITC-containing ROIs are also mislabeled as benign in the ground truth. These are examples of improper true positives. **6B**. A 40x FOV with model predictions, annotations, and immunostain. This FOV is the upper left quadrant of the above 20x FOV. All false negatives in this FOV are proper.

## Discussion

To the best of our knowledge, this is the first paper that attempts to understand how a deep learning model performs on the cell and tissue types found in the lymph node biopsy. Similar to how early studies in immunohistochemistry identified biotin, peroxidase, and collagen as sources of non-specific binding, we have identified the subclasses of tissue which the LYNA models have difficulty distinguishing from cancer. This is significant because models for medical imaging commonly suffer from hidden stratification (19). For instance, the overall performance of a cancer detection algorithm may be high, but its performance on an uncommon cancer subclass may be extremely poor. This discrepancy in performance would be extremely important for pathologists to be aware of but may go unnoticed if the domain-relevant distinctions were not captured in testing.

We find that the LYNA models performed best on the blood and fat tissue subclasses, likely due to their visual dissimilarity from cancer. The ‘fat’ subclass also demonstrates the importance of testing on other types of tissue. The models’ high performance on fat tissue (approx. accuracy 0.99) and its prevalence within our test set (approx. 70% of ROIs) greatly inflated the overall performance metrics. The models struggled most with the traditionally difficult subclasses of histiocytes and germinal centers, but also performed poorly on the lymphocytes, capsule, and sinus tissue. In the case of the sinus subclass, the models’ performance significantly increased with magnification. We hypothesize that this was because the sinus subclass ROIs came to contain fewer resident histiocytes and more sinus endothelial cells as magnification increased. Conversely, the models’ performance on metastatic breast cancer decreased with magnification. Variable performance among models operating on different magnifications may present an issue for the usage of AI models for decision support. At the very least, it is critical for operational users to understand the limitations of the model at a given resolution. Figure 5 presents an example of the LYNA models’ variable performance across 20x and 40x magnification FOVs.

It is important to note that these results obtained at the ROI level do not have a good point of comparison within the literature; they are not directly comparable to results found for models which detect cancer on a coarser resolution, such as on WSIs. On the other hand, the FOV-level task we performed is directly comparable to the task in Chen *et al*. (2019). On this task, we obtained values of 0.94 (95% CI, 0.920-0.951) and 0.96 (95% CI, 0.953-0.964) AUC at 10x and 20x magnifications respectively, which aligns well with the values of 0.92 and 0.97 AUC which they reported (17). The LYNA models on our FOV-level task obtained sensitivities and NPVs which were comparable or higher than the values obtained at the ROI level. These metrics indicate that the models’ ability to correctly classify actual cancer is similar at both levels and consistent with previous performance.

In addition, the models’ performance on some types of friable tumors in the out-of-domain test set, such as high-grade carcinoma, indicates the potential for the LYNA models to detect more types of clinically relevant cancer. The comparatively lower positive predictive value of the models on endometrial carcinoma and germ cell tumor (0.58 and 0.67 respectively) can partially be attributed to the existence of benign tissue within the ROIs: entrapped benign endometrium in the case of endometrial carcinoma and desmoplastic stroma in the case of the mixed germ cell tumor. The low positive predictive value on borderline tumors compared to other cancers is consistent with the findings of Liu *et al*. (2019), which described the LYNA model’s tumor detection response to large, disordered nuclei, but not smaller, non-malignant nuclei (18). After testing, we examined ROIs where model classifications diverged from ground truth to understand common sources of error within both model performance and ground truth annotations. We classified each of these errors as *proper* or *improper*, as defined in the Materials and Methods.

We found that improper false negative errors were most commonly due to the over-labeling of cancer by the annotating pathologist. As demonstrated in Figure 4C, over-labeling occurred when the annotations were drawn at a low magnification around regions of tissue which contained large amounts of cancer. These imprecise annotations produced ROIs which were improperly labeled as cancer, especially at the higher magnifications of 20x and 40x. This over-labeling frequently occurred for large, complex groups of tumor cells or clusters of isolated tumor cells. These regions are widely recognized to be challenging for pathologists and have been shown to be an area where AI models may be helpful in clinical practice (16). In addition, we hypothesize that the models’ decreasing performance on breast cancer at higher magnifications may be partially attributable to improper false negatives at the boundary. This is caused by the fact that cancerous regions were often annotated with a small “buffer” around the cancer. This imprecision in our tumor annotations would penalize performance at higher magnifications, since smaller ROI sizes would cause more of these buffer regions to become improper cancer ROIs.

Our examination also found ITCs as a source of both model and ground truth error. Although we did not separate ITC-containing ROIs into a separate subclass of cancer, we found that the models had difficulty in detecting these ROIs, especially at lower magnifications. This is consistent with the performance of other successful deep learning models on ITCs; the best model within the CAMELYON17 challenge was only able to identify ITCs with an 11.4% accuracy (20). Our re-review also indicated that many ITC-containing ROIs were improperly labeled as ground truth benign at the 10x and 20x magnifications due to their vanishingly small size. Despite both classes of errors, performance on ITCs did not significantly affect average performance metrics due to their relatively low occurrence and small area. As such, they present an example of hidden stratification; if performance over ITCs is not separately considered from performance over other types of cancer, it can easily be lost in aggregate metrics. It is important to note that ITCs are also difficult for pathologists to detect and do not affect treatment decisions today.

This detailed level of analysis that we performed, such as ROI-level performance across subclasses, was only enabled by the extremely detailed level of annotation that digital pathology allows. It is difficult to imagine building and annotating a dataset with 48,477 FOVs captured directly from traditional photomicrographs, especially a dataset that annotated each tissue subclass. Digital labeling of WSIs allowed for the reduction of manual labor, continuity of annotation between FOVs, and the ability to reuse labels between magnifications. Even with all of this, our dataset consists of only 40 WSIs, essentially the bare minimum needed for clinical validation.

Future assessments of decision support algorithms may consider other metrics for quantifying error. Within this paper, we treated the classifications of each ROI as independent and equal, disregarding the geometry of the FOVs and their organization within the test set. A disadvantage of this approach is that it ignores the contextual importance of a classification. For example, ten false negatives at the border of a macrometastasis which was mostly detected may be less consequential than ten false negatives which constitute a micrometastasis. In practice, the former would result in a slightly smaller-than-correct outline on the ARM, while the latter would result in an entirely missed tumor. An alternative metric which would differentiate the above cases is the intersection over union value.

Overall, this paper provides a template for the analysis of an ARM-optimized model’s performance. Examining metrics at the ROI level (as opposed to at an FOV or WSI level) allows us to understand the model’s suitability for usage in the decision support capacity. Examining model performance across tissue subclasses uncovers previously hidden nuances about the models’ performance characteristics. Finally, the analysis of ground truth using the proper/improper distinction provides the ability to understand annotator weaknesses and quantify uncertain ground truth. We hope that this paper will be useful in the validation of other AI models that are integrated onto the ARM platform to improve the microscopy workflow for the diagnosis of cancer or other diseases.

### Limitations & Future Work

This study has some important limitations.

First, all WSIs used within the testing set were sourced from a single medical center and labeled by a single pathologist. The labeling pathologist was aided with each slide’s immunostain and was constrained to four hours of labeling time per WSI. This limitation led to the lack of a second opinion or additional resources to decide difficult diagnoses, as well as the labeling at a lower than optimal resolution.

Second, this study used WSIs to emulate the magnification and pixel size of microscopic FOVs instead of capturing the photos directly from a microscope. While this was necessitated by the required level of annotation, the use of digitized FOVs limits the generalizability of this study to augmented reality. In particular, the FOVs used within this study differ from microscopic FOVs in their lack of traditional microscope artifacts of vignetting, blur, and fisheye. Additionally, the process of obtaining a ground truth annotation from the WSI magnification resulted in some inaccurate ground truth ROIs due to thresholding.

Third, this study does not directly provide evidence to the efficacy of the LYNA models on the ARM as a decision support system. In order to establish such claims, one must consider the ARM-pathologist team, rather than the model, as the primary object of study. This may be analogous to the work in Steiner *et al*. (2018), which considered the performance of the model-assisted pathologist, rather than just the model itself (16).

Future work should include ground truth annotations by multiple pathologists, FOV data gathered from ARMs fielded in laboratories, prospective trials, and similar analysis on other deep learning models that have been adapted for the ARM platform, such as models for Gleason grading, mitosis counting, and cervical dysplasia grading.

## Data Availability

The datasets used and analyzed in this study are available from the corresponding author upon reasonable request.

## Acknowledgements

The authors wish to acknowledge the following individuals for their help in this study: Jeremy Lai, Brian Nock, Sofia Blasini, Tony Carnevale, Cameron Chen PhD, Fraser Tan PhD, Erik Seetao, James Wren, Josh Pomorski, Briana Rivas, Nathaniel Higgins, Pierre Samanni, Danielle Long-Hyland Capt USAF, Hassan Tetteh MD CAPT USN, Brian Woolley PhD LTC USAF, Anthony DiGregorio, Laura Mariano, Arash Mohtashamian MD CAPT USN, Carrie Robinson MD LCDR USN, Brandon Peterson MD LCDR USN, Jen Lombardo, Jon Elliott, and Jessica Rajkowski.

