## Supplementary Materials for "Independent assessment of a deep learning system for lymph node metastasis detection on the Augmented Reality Microscope"

### Model details

The LYNA models are built upon Inception v3. The models use a sliding window approach to evaluate each ROI. This allows for the inference of each ROI to take context from the neighboring regions for their predictions, which emulates the workflow of a pathologist. This is done by using a computationally-optimized fully convolutional network which reduces the recomputations of overlapping areas between ROIs (Chen *et al.*, 2019).

The FOV is a 1800x1800 RGB pixel image which is fed into the model. Before inference, 4 pixels are trimmed on each side. The model performs inference on a region of 1792x1792 pixels. This 1792x1792 area is divided into a 14x14 grid of 128x128 pixel ROIs (14*128 = 1792). The inference performed on each ROI within the model also contains the surrounding context. The input ROI has a buffer area of ~392 pixels on each side to have a total area of 911 x 911 pixels. Thus, if we include the buffer on the edge of ROIs, we have a total region of 2575x2575 (1792 + 392 + 391 = 2575) which the model ultimately interacts with. This surrounding buffer is outside of the ARM's FOV (Chen *et al.*, 2019).

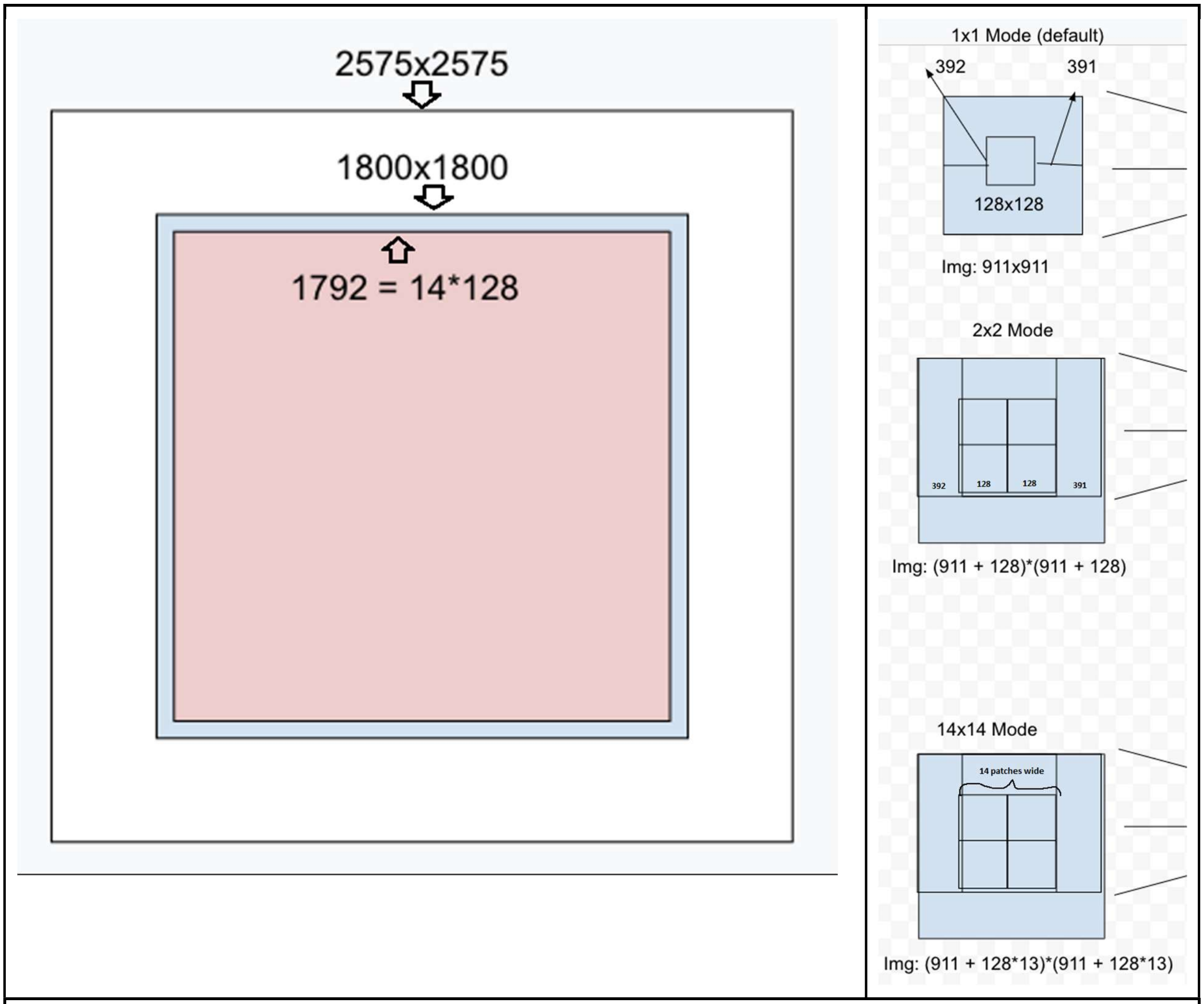


**Supplementary Figure 1. Model input sizes**
1A. Comparison of FOV input sizes. The blue square indicates the size of the initial FOV. The red square indicates FOV size after trimming. The white square indicates the full area that the model interacts with, including the buffer.
1B. ROI size and buffers. The 1x1, 2x2, and 14x14 modes indicate the total pixel size of inference for various numbers of patches at a time, as well as the areas of overlap. The model operates on the 14x14 mode shown within the figure. Thus, the model will ultimately output 14*14 = 196 inferences for a single FOV.

### Synthetic vs Microscopic FOVs

Within microscopic FOVs, vignetting and image-warping near the outside of the FOV make diagnosis in these regions infeasible for both models and pathologists. For ROIs within these regions, the LYNA model's performance is significantly degraded. Thus, in the actual evaluation of microscopic FOVs, the performance of the LYNA models on ROIs which are subject to too much distortion would be excluded from the testing data. The use of synthetic FOVs derived from whole slide images allows for all ROIs within the FOV to be used for testing.

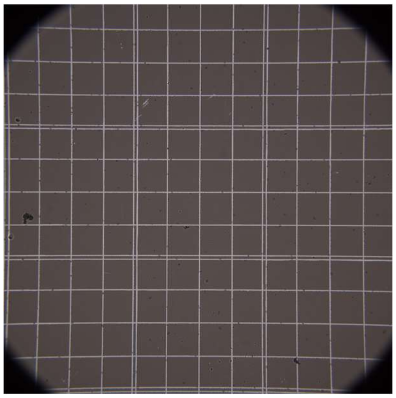

Camera FOV

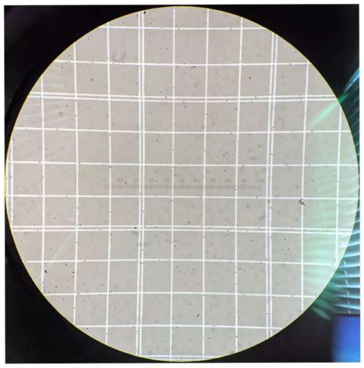

Ocular FOV

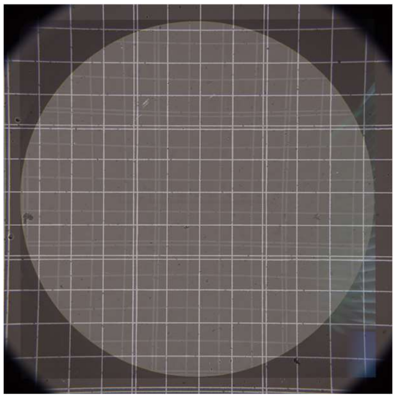

Camera and Ocular FOV overlaid

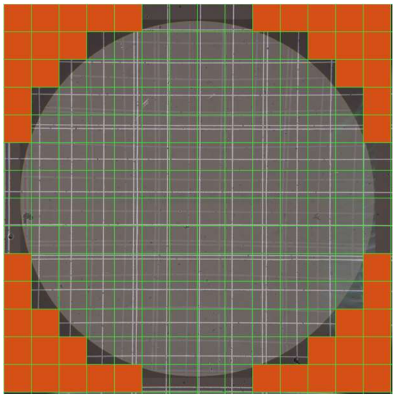

Maximum visible patches, based on minimally omitable ROI patches

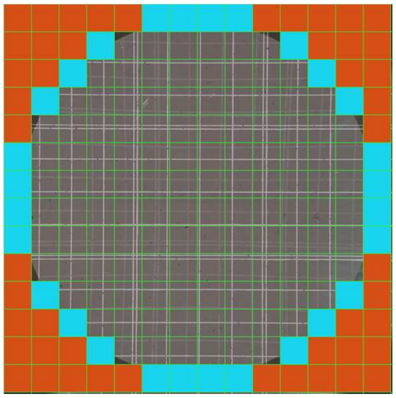

Moderately visible patches based on moderately omitable ROI patches

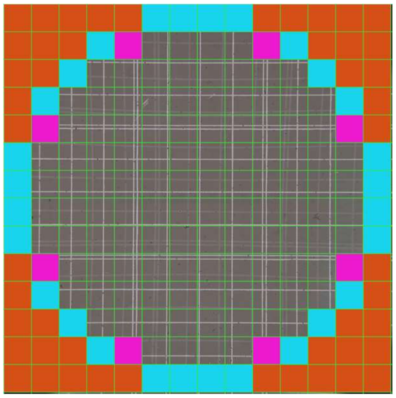

Minimum visible patches based on maximally omitable ROI patches

**Supplementary Figure 2. Visibility of various FOV regions on ocular and camera FOVs**
Within microscopic FOVs, the outermost ROIs are sufficiently distorted to impact model performance and be omitted from testing.

### Ground truth labeling procedure

The FOV in Supplementary Figure 3 gives insight into the way in which pixel-level annotations are turned into classifications for each ROI. An ROI which has 10% or more of its area annotated as cancer is given the “cancer” ground truth label. Otherwise, if more than 50% of its area is annotated, it is given the label of the most common benign subclass within its area. If less than 50% of its area is annotated, it is not given a ground truth annotation and excluded from the testing set.

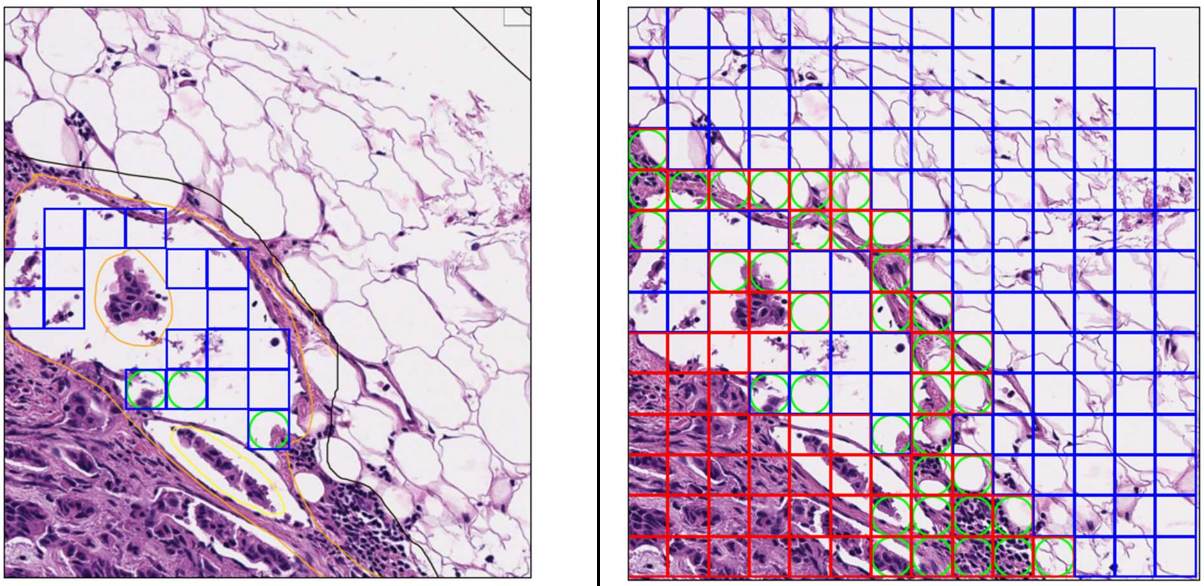

**Supplementary Figure 3. 40x Sinus FOV annotation example**
3A. FOV annotation overlaid with sinus ground truth. Sinus is annotated in orange and ROIs which are ground truth "Sinus" and overlaid.
3B. FOV with full ground truth overlay, including other tissue types. Note false negatives at the border compared to the annotation outline

Further results over primary test set

**Supplementary Table 1. ROI and FOV level results summary**
95% confidence interval in brackets calculated using 1000 bootstrap iterations.

| Model | Mode | AUC | Accuracy | Sensitivity | Specificity | PPV | NPV |
|---|---|---|---|---|---|---|---|
| LYNA 10x | ROI | 0.981 [0.9810, 0.9821] | 0.938 [0.9368, 0.9383] | 0.930 [0.9277, 0.9327] | 0.938 [0.9375, 0.9390] | 0.604 [0.6006, 0.6088] | 0.993 [0.9923, 0.9928] |
| | FOV | 0.938 [0.9204, 0.9507] | 0.746 [0.7325, 0.7621] | 0.939 [0.9214, 0.9631] | 0.716 [0.7009, 0.7330] | 0.342 [0.3164, 0.3667] | 0.987 [0.9826, 0.9919] |
| LYNA 20x | ROI | 0.978 [0.9779, 0.9785] | 0.939 [0.9381, 0.9389] | 0.897 [0.8953, 0.8987] | 0.943 [0.9421, 0.9429] | 0.601 [0.6984, 0.6034] | 0.990 [0.9894, 0.9898] |
| | FOV | 0.959 [0.9534, 0.9644] | 0.792 [0.7815, 0.7986] | 0.956 [0.9413, 0.9669] | 0.771 [0.7603, 0.7785] | 0.347 [0.3298, 0.3656] | 0.993 [0.9901, 0.9948] |
| LYNA 40x | ROI | 0.976 [0.9757, 0.9761] | 0.947 [0.9470, 0.9474] | 0.878 [0.8768, 0.8787] | 0.954 [0.9536, 0.9540] | 0.641 [0.6403, 0.6429] | 0.988 [0.9880, 0.9881] |
| | FOV | 0.948 [0.9453, 0.9515] | 0.823 [0.8192, 0.8261] | 0.948 [0.9422, 0.9547] | 0.809 [0.8051, 0.8125] | 0.354 [0.3453, 0.3658] | 0.993 [0.9920, 0.9938] |

| **Supplementary Table 2. Results per subclass per magnification**<br>95% confidence interval in brackets calculated using 1000 bootstrap iterations. | | | | | | | |
|---|---|---|---|---|---|---|---|
| | | 10x | | 20x | | 40x | |
| Class | Subclass | Accuracy | % of ROIs | Accuracy | % of ROIs | Accuracy | % of ROIs |
| Cancer | BrCA | 0.930 [0.9278, 0.9337] | 9.19% | 0.897 [0.8955, 0.8986] | 8.80% | 0.878 [0.8769, 0.8787] | 8.61% |
| Immune cells | Histiocytes | 0.547 [0.5328, 0.5677] | 0.90% | 0.532 [0.5241, 0.5431] | 0.98% | 0.564 [0.5600, 0.5683] | 0.99% |
| | GC | 0.675 [0.6119, 0.7351] | 0.08% | 0.758 [0.7375, 0.7820] | 0.09% | 0.683 [0.6694, 0.6956] | 0.09% |
| | Mantle | 0.869 [0.8272, 0.9162] | 0.05% | 0.943 [0.9285, 0.9603] | 0.06% | 0.932 [0.9227, 0.9394] | 0.06% |
| | Lymphocytes | 0.777 [0.7731, 0.7801] | 17.81% | 0.778 [0.7762, 0.7793] | 17.71% | 0.821 [0.8203, 0.8218] | 17.74% |
| Connective Tissue | Sinus | 0.437 [0.3544, 0.4951] | 0.06% | 0.615 [0.5904, 0.6451] | 0.06% | 0.783 [0.7717, 0.7955] | 0.06% |
| | Capsule | 0.746 [0.7354, 0.7549] | 1.28% | 0.757 [0.7518, 0.7623] | 1.54% | 0.827 [0.8203, 0.8218] | 1.59% |
| | Nerve | 0.914 [0.8143, 0.9714] | 0.02% | 0.839 [0.8017, 0.8822] | 0.02% | 0.948 [0.9383, 0.9582] | 0.02% |
| | Artery | 0.900 [0.8793, 0.9212] | 0.21% | 0.923 [0.9139, 0.9398] | 0.24% | 0.956 [0.9512, 0.9587] | 0.25% |
| | Vein | 0.930 [0.9191, 0.9393] | 0.66% | 0.918 [0.9134, 0.9232] | 0.72% | 0.954 [0.9521, 0.9555] | 0.79% |
| | Blood | 0.998 [0.9961, 0.9988] | 0.93% | 0.997 [0.9961, 0.9981] | 0.94% | 0.997 [0.9961, 0.9973] | 0.95% |
| | Fat | 0.989 [0.9883, 0.9893] | 68.81% | 0.995 [0.9950, 0.9953] | 68.82% | 0.996 [0.9963, 0.9964] | 68.89% |

| Supplementary Table 3. Number of FOVs and ROIs per magnification | | |
|---|---|---|
| Magnification | Total number of FOVs | Total number of ROIs |
| 10x | 2,905 | 358,285 |
| 20x | 10,018 | 1,448,284 |
| 40x | 35,554 | 5,802,458 |
| Total | 48,477 | 7,609,027 |

| Supplementary Table 4. ROI level confusion matrix, primary and without fat datasets, per magnification | | | | | |
|---|---|---|---|---|---|
| Model | Test Set | True Positive | True Negative | False Positive | False Negative |
| LYNA 10x | Primary | 30637 | 305273 | 20083 | 2292 |
| | No fat | 30637 (100%) | 61488 (20.1%) | 17337 (86.3%) | 2292 (100%) |
| LYNA 20x | Primary | 114347 | 1244949 | 75915 | 13073 |
| | No fat | 114347 (100%) | 253074 (20.3%) | 71057 (93.6%) | 13073 (100%) |
| LYNA 40x | Primary | 438542 | 5057601 | 245248 | 61067 |
| | No fat | 438542 (100%) | 1075065 (21.3%) | 230728 (94.1%) | 61067 (100%) |

| Supplementary Table 5. FOV level confusion matrix per magnification | | | | |
|---|---|---|---|---|
| Model | True Positive | True Negative | False Positive | False Negative |
| LYNA 10x | 371 | 1797 | 713 | 24 |
| LYNA 20x | 1084 | 6848 | 2036 | 50 |
| LYNA 40x | 3358 | 25906 | 6105 | 185 |

**Supplementary Table 6. Cancer ROIs per cancer WSI**
Slides 1-15 contain macrometastases. Slides 16-20 contain only micrometastases and are indicated in bold.

| slide # | 10x | | 20x | | 40x | |
|---|---|---|---|---|---|---|
| | # of ROIs | % of cancer | # of ROIs | % of cancer | # of ROIs | % of cancer |
| 1 | 13,599 | 41.30% | 53,154 | 41.73% | 210,050 | 42.05% |
| 2 | 4,755 | 14.44% | 18,532 | 14.55% | 73,192 | 14.65% |
| 3 | 4,445 | 13.50% | 17,331 | 13.61% | 68,451 | 13.70% |
| 4 | 1,998 | 6.07% | 7,752 | 6.09% | 30,509 | 6.11% |
| 5 | 1,893 | 5.75% | 7,349 | 5.77% | 28,904 | 5.79% |
| 6 | 1,729 | 5.25% | 6,741 | 5.29% | 26,551 | 5.32% |
| 7 | 1,701 | 5.17% | 6,535 | 5.13% | 25,526 | 5.11% |
| 8 | 762 | 2.31% | 2,592 | 2.03% | 9,401 | 1.88% |
| 9 | 414 | 1.26% | 1,488 | 1.17% | 5,154 | 1.03% |
| 10 | 339 | 1.03% | 1,249 | 0.98% | 4,586 | 0.92% |
| 11 | 302 | 0.92% | 986 | 0.77% | 3,429 | 0.69% |
| 12 | 242 | 0.73% | 907 | 0.71% | 3,501 | 0.70% |
| 13 | 226 | 0.69% | 846 | 0.66% | 3,239 | 0.65% |
| 14 | 218 | 0.66% | 862 | 0.68% | 3,175 | 0.64% |
| **15** | 57 | 0.17% | 199 | 0.16% | 722 | 0.14% |
| **16** | 151 | 0.46% | 530 | 0.42% | 1,926 | 0.39% |
| **17** | 39 | 0.12% | 128 | 0.10% | 476 | 0.10% |
| **18** | 25 | 0.08% | 84 | 0.07% | 297 | 0.06% |
| **19** | 22 | 0.07% | 74 | 0.06% | 265 | 0.05% |
| **20** | 12 | 0.04% | 38 | 0.03% | 116 | 0.02% |

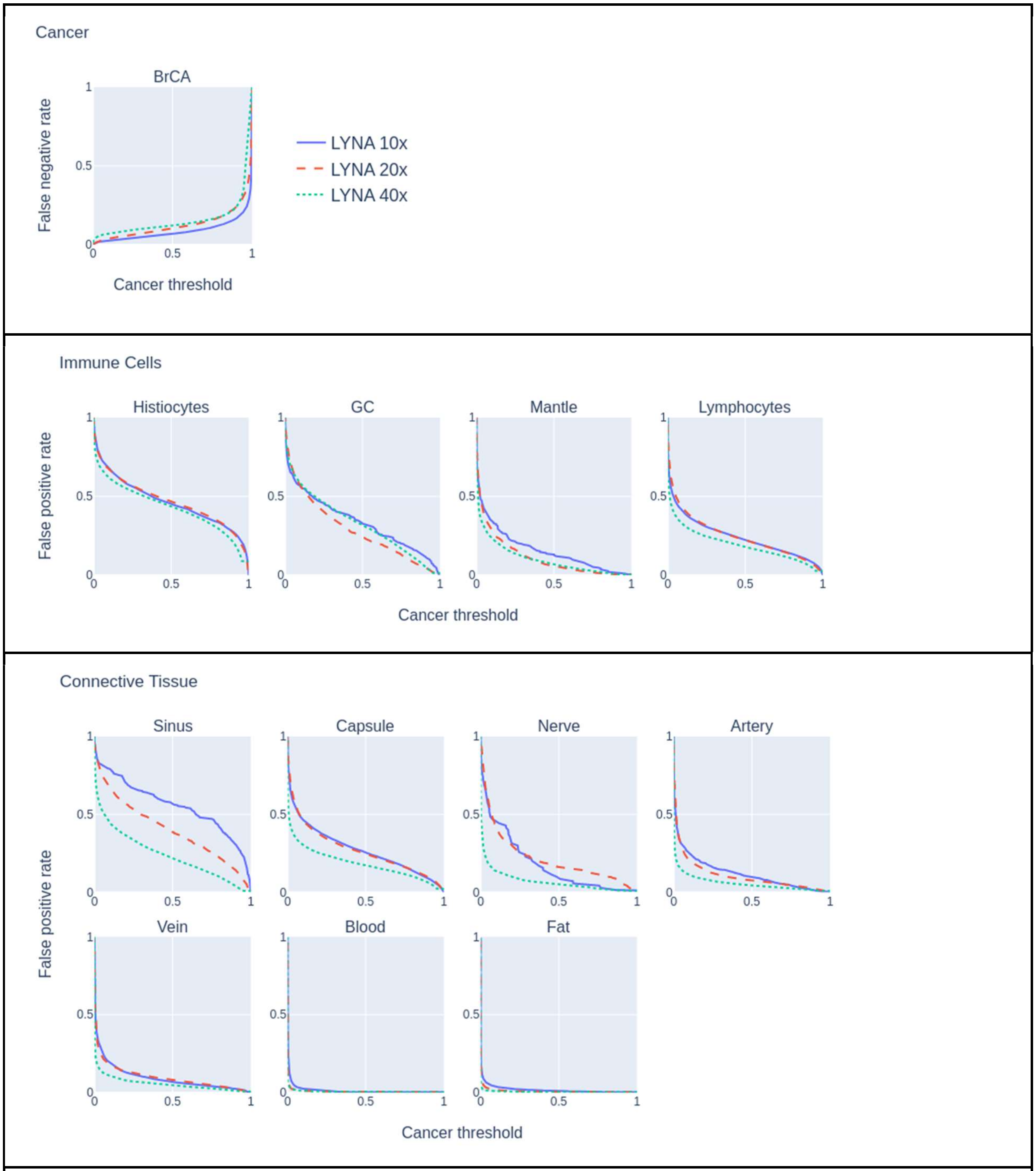


**Supplementary Figure 4. Cancer threshold vs error rate per subclass, model, and magnification**
4A. The tradeoff between cancer threshold (0.0 to 1.0) and false negative rate for each LYNA model on the breast cancer subclass.
4B. The tradeoff between cancer threshold (0.0 to 1.0) and false positive rate for each LYNA model across each immune cell subclass.
4C. The tradeoff between cancer threshold (0.0 to 1.0) and false positive rate for each LYNA model across each connective tissue subclass.

### Fat subclass ablation study

**Supplementary Table 7. ROI-level results summary, removing fat**
Primary test set results presented for comparison

| Model | Test Set | AUC | Accuracy | Sensitivity | Specificity | PPV | NPV |
|---|---|---|---|---|---|---|---|
| LYNA 10x | No fat | 0.9408 | 0.8244 | 0.9304 | 0.7801 | 0.6386 | 0.9641 |
| | Primary | 0.9816 | 0.9375 | 0.9304 | 0.9383 | 0.6040 | 0.9925 |
| LYNA 20x | No fat | 0.9231 | 0.8137 | 0.8974 | 0.7808 | 0.6167 | 0.9509 |
| | Primary | 0.9783 | 0.9386 | 0.8974 | 0.9425 | 0.6010 | 0.9896 |
| LYNA 40x | No fat | 0.9204 | 0.8384 | 0.8778 | 0.8233 | 0.6553 | 0.9463 |
| | Primary | 0.9759 | 0.9472 | 0.8778 | 0.9538 | 0.6413 | 0.9881 |

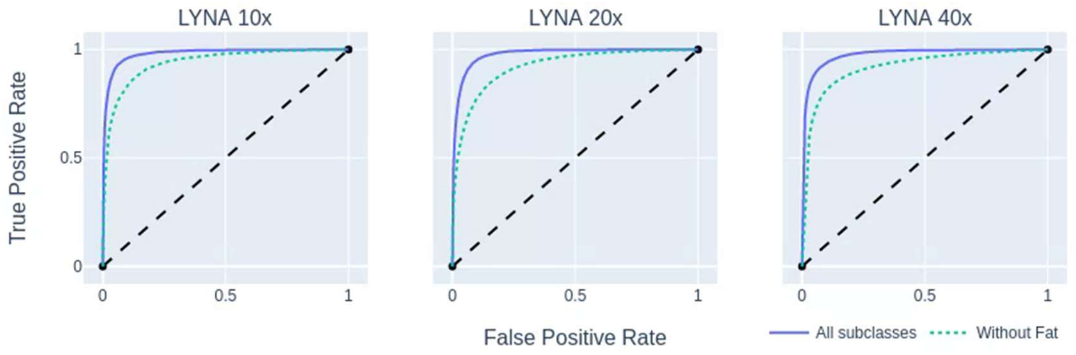


**Supplementary Figure 5. ROI level ROC curves with and without fat per magnification**
Comparison of the ROC curves of each model on the standard test set and the test set with the "fat" subclass removed.

### Out-of-domain results

| **Supplementary Table 8. ROI level accuracy, OOD test set, 10x magnification** | | |
|---|---|---|
| Tissue type | Accuracy | # of ROIs |
| High grade carcinoma | 0.949 | 5894 |
| Papillary thyroid cancer | 0.902 | 3663 |
| Papillary urothelial carcinoma | 0.728 | 1540 |
| Endometrial carcinoma | 0.579 | 4582 |
| Embryonal carcinoma | 0.672 | 8391 |
| Serous borderline tumor | 0.093 | 12251 |

### Errors per WSI

In order to better understand the types of errors that are occurring, we analyzed the distribution of false negative ROIs among the 20 WSIs containing metastatic breast cancer within our testing dataset. A summary graph of the distribution of errors across slides can be found in Supplementary Figure 6.

For each WSI, the percentage of the total cancer ROIs remains stable across the resolutions. This is as expected because the ROIs derive their cancer labels from the same ground truth annotation. However, for a given WSI, there is inconsistency for the percentage of its cancer ROIs that are in error across resolutions. This inconsistency indicates different levels of performance between the LYNA models of varying resolutions on FOVs of the same underlying area. There is no model that is clearly outperforming the others; the relative levels of performance greatly varies from slide to slide. Within the WSIs with macrometastases, the slide with the most errors at each magnification is Supplementary Figure 7. This WSI had sections of its immunostain damaged, and thus, was partially labeled without the help of an immunostain. We hypothesize that this contributed greatly to the amount of disagreement found in this slide. Within the WSIs with only micrometastases, performance of the 10x LYNA model is significantly worse than the higher resolutions, which we believe is partially attributable to the low sample size of ground truth cancer ROIs at low magnifications.

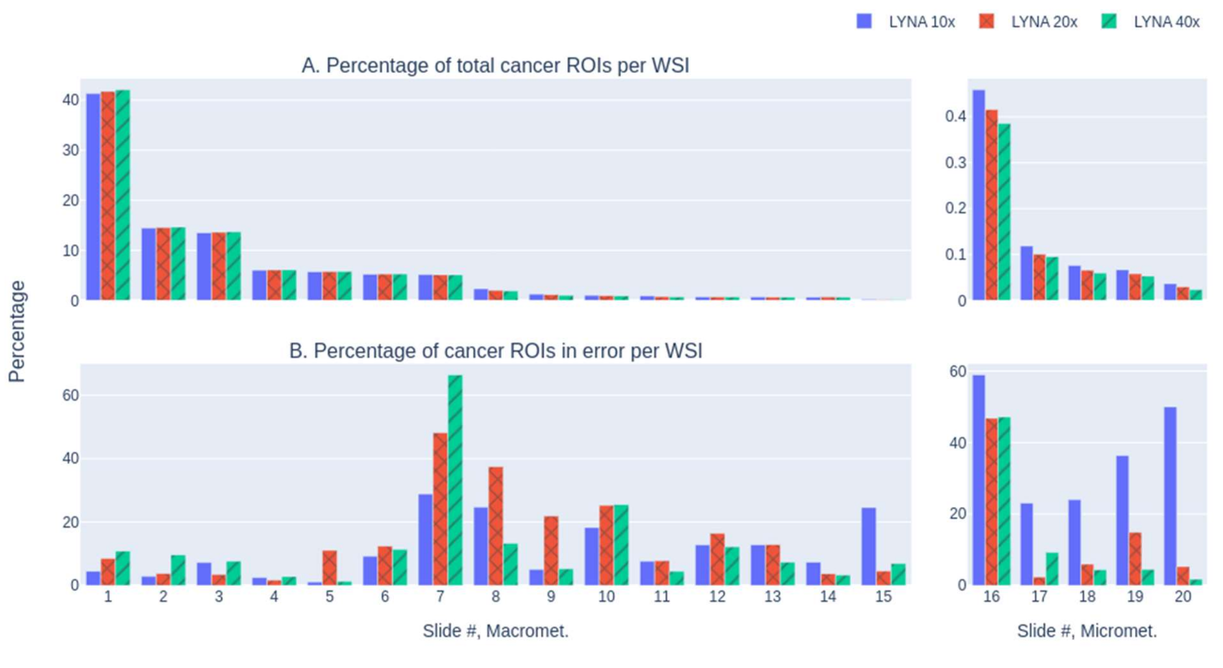


**Supplementary Figure 6. Summary graphs of cancer ROIs and false negative ROIs per WSI**
6A. For each magnification and each WSI, the percentage of the total cancer ROIs (for that magnification) contained in that WSI. Macromets and micromets plotted separately.
6B. For each magnification and each WSI, the percentage of cancer ROIs that are false negative errors in that WSI. Macromets and micromets plotted separately.

### Examination of errors

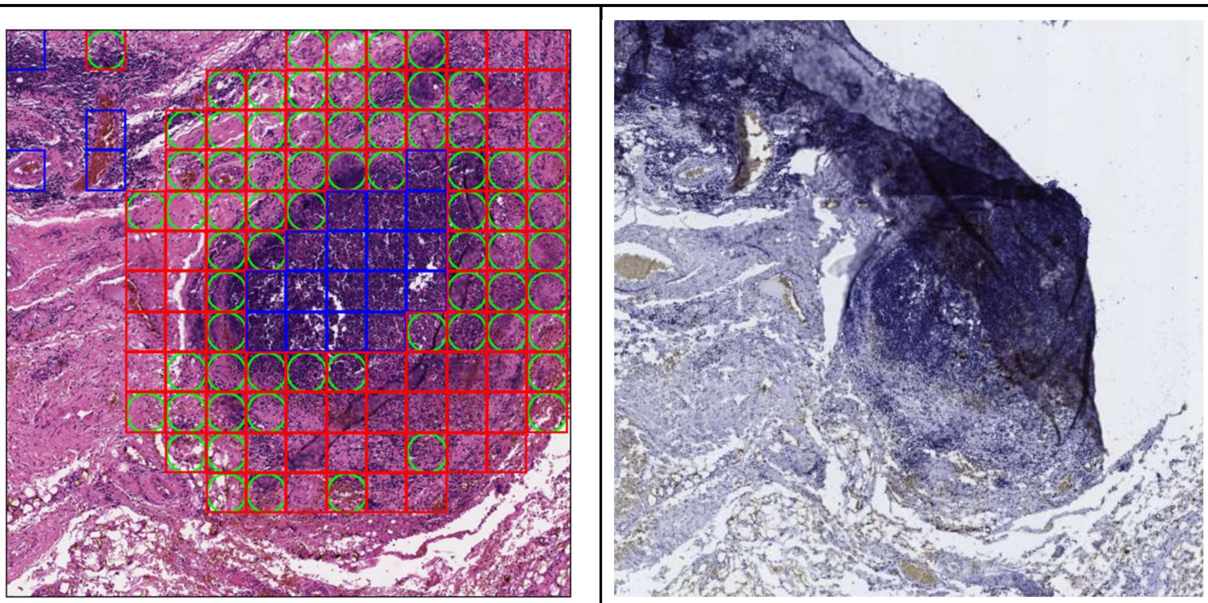

**Supplementary Figure 7. 10x FOV, indeterminate false negatives**
7A. 10x magnification FOV with model predictions
7B. 10x magnification FOV immunostain

This FOV has a large number of false negatives in its central region. During the restaining procedure, the tissue was disrupted and folded over upon itself, resulting in the above damaged immunostain. The FOV in consideration fell within this damaged region, so the annotating pathologist labeled this FOV without immunostain support. Given the lack of immunostain, we classified the false negatives in the above FOV as having indeterminate disagreement.

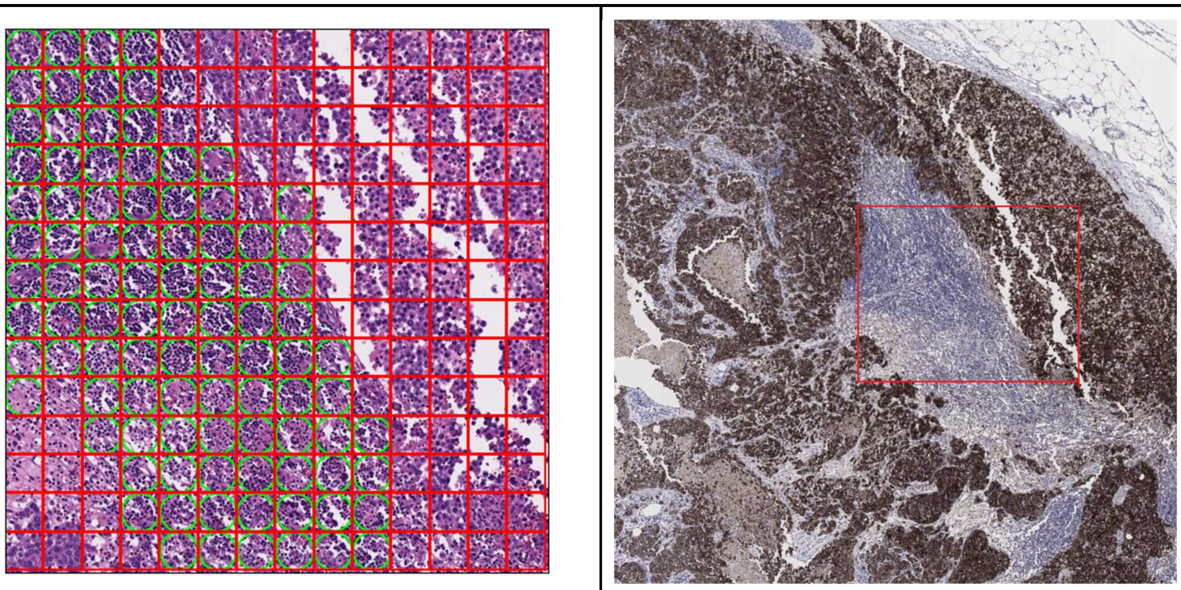

**Supplementary Figure 8. 20x FOV, improper false negatives**
8A. 20x magnification FOV with model predictions
8B. Slide immunostain, with box indicating corresponding FOV region

On review, we concluded that most of the false negative ROIs within this FOV are the result of over-labelling of cancer, again driven by time constraint. Thus, the majority of the false negative ROIs within this FOV are improper false negatives.

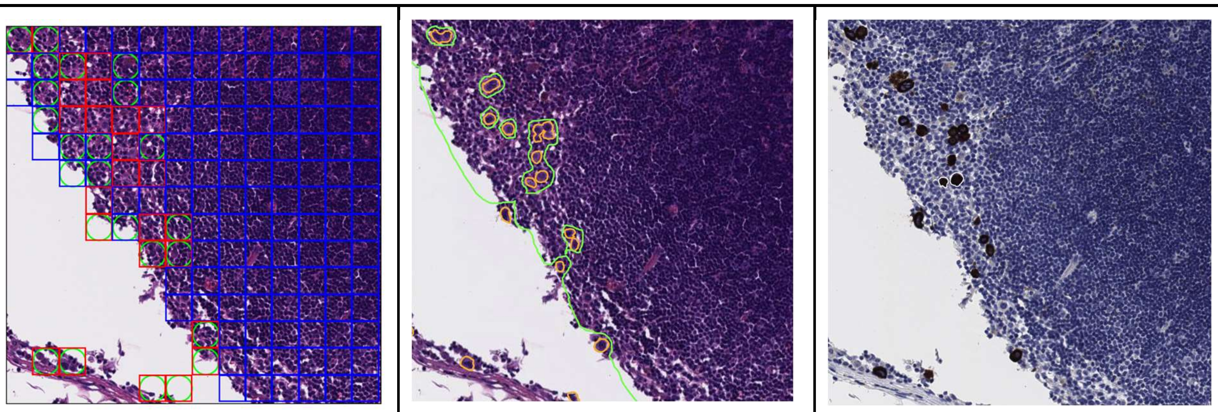

**Supplementary Figure 9. 40x FOV, proper & improper false negatives**
9A. 40x magnification FOV with model predictions
9B. 40x magnification FOV with annotation
9C.40x magnification FOV immunostain

Annotating the ITCs in this FOV requires several complex boundary decisions and those boundaries interact with the ground truth assignment in various ways.
On detailed review, we conclude the following: the false negatives along the bottom of the FOV are proper false negatives. Some false negatives within the slide are improper, due to boundary issues with ground truth determination. These false negatives involving ITCs compose a very small minority of the errors.

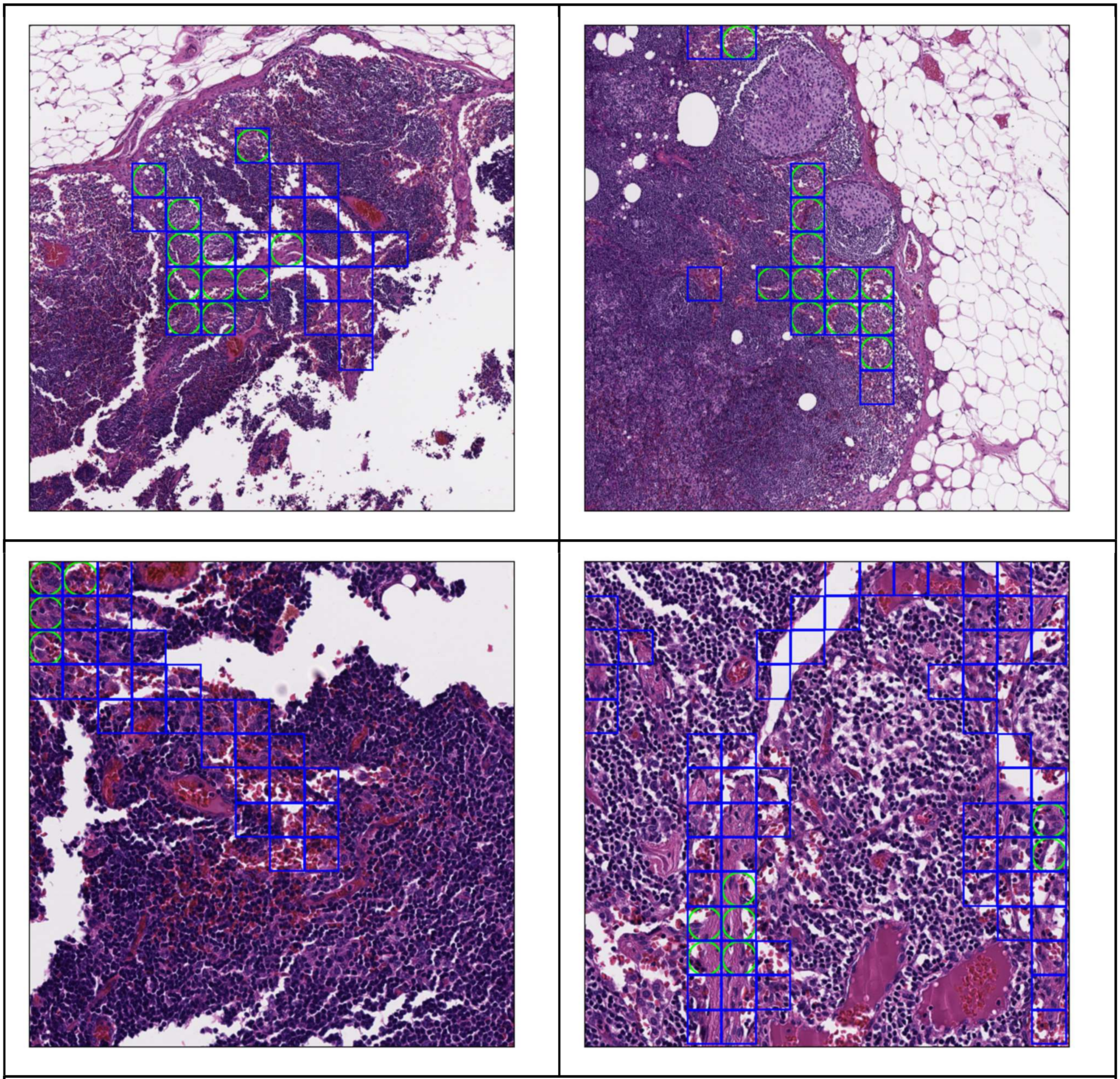

**Supplementary Figure 10. 10x & 40x FOVs, proper false positives**
10A-B. 10x FOV with model predictions on sinus subclass only
10C-D. 40x FOV with model predictions on sinus subclass only
In the above FOVs, only sinus ROIs are bounded with boxes. Since sinus is a benign subclass, all ROIs indicated represent ground truth benign ROI. Within the 10x FOVs, the false positives are proper on review. Many of the sinus ROIs in the 10x FOVs have a significant proportion of histiocytes within their area.
Within the 40x FOVs, the false positives are also proper, but there are far fewer of them. The higher resolution at 40x FOVs makes the ground truth subclass for a given ROI more precise, which allows sinus ROIs to generally contain a lower proportion of other cells such as histiocytes.
